# The analysis of risk factors associated with multimorbidity of anaemia, malaria, and malnutrition among children aged 6- 59 months in Nigeria

**DOI:** 10.1101/2023.04.10.23288389

**Authors:** Phillips Edomwonyi Obasohan, Stephen J. Walters, Richard Jacques, Khaled Khatab

## Abstract

In the last ten years multimorbidity in children under the age of five years has becoming an emerging health issue in developing countries. The absence of a proper understanding of the causes, risk factors, and prevention of these new health disorders (multimorbidity) in children is a significant cause for concern, if the sustainable development goal 3 of ensuring healthy lives and the promotion of well-being for all especially in the associated aim of ending preventable deaths of new-borns and children must be achieved by 2030. In the past, most studies conducted in Nigeria and some other least developed nations of the world focused on these multiple diseases by employing conventional analytical techniques to examine them separately as distinct disease entities. But the study of multimorbidity of anaemia, malaria, and malnutrition has not been done, especially in children. This study aims to investigate the multiple overlaps in the impact of individual and contextual variables on the prevalence of multimorbidity among children aged 6 to 59 months in Nigeria. The study used two nationally representative cross-sectional surveys, the 2018 Nigeria Demographic and Health Survey and the 2018 National Human Development Report.

A series of multilevel mixed effect ordered logistic regression models were used to investigate the associations between child/parent/household variables (at level 1), community-related variables (at level 2) and area-related variables (at level 3), and the multimorbidity outcome (no disease, one disease only, two or more diseases). The interaction effects between child’s sex, age, and household wealth quintiles and the outcome while accounting for some covariates in the model were also investigated. The result shows that 48.3% (4,917/10,184) of the sample of children aged 6-59 months cohabit with two or more of the disease outcomes. Child’s sex, age, maternal education, mother’s anaemia status. household wealth quintiles, the proportion of community wealth status, states with high human development index, region, and place of residence, were among the significant predictors of MAMM (p<0.05). There was a significant interaction effect between a child’s age and wealth status when some other covariates were accounted for. The prevalence of MAMM observed in the sample is large, with almost half of the children living with two or more of the diseases at the time of the survey and several potentially modifiable risk factors have been identified. If suitable actions are not urgently taken, Nigeria’s ability to actualise the SDG 3 will be in grave danger. Therefore, possible actions to ameliorate this problem include developing and implementing a suitable policy that will pave the way for integrated care models to be developed.

## Introduction

Childhood mortality and morbidity rates are still very high, especially in Low-and Medium-Income Countries (LMIC) and these have resulted in a severe public health burden [1]. According to the World Health Organization (WHO) and the Centres for Disease Control and Prevention (CDC), about 25% of the world’s population is anaemic, with expectant mothers and children under the age of five years being the most vulnerable [2–4], but since 2016, the prevalence of anaemia has increased globally by more than 40% annually [5]. Similarly, over the last twenty years, malaria has remained a primary public health concern[6], with over 300 million cases reported in 2018 [7]. It has remained a leading cause of morbidity and mortality with LMICs, especially Sub-Saharan Africa (SSA), contribute more than 80% of the global malaria burden [8,9]. Though a considerable global decline has been noticed in childhood stunting, over 150 million, 50 million and 38 million children remain stunted, wasted and overweight, respectively [10]. However, in 2018, there were more than 40 million overweight children under the age of five years, which was contrary to expectations and in keeping with a global target on malnutrition to keep the rate of childhood obesity constant. [11], indicating a gradual global rise in overweight children. It becomes more worrisome when children simultaneously co-inhabit two or more of these disease conditions.

There is evidence in research that anaemia, malaria, and malnutrition interrelate, resulting in adverse health outcomes and mortality, especially in children under-five years. In a recent study conducted on SSA, malnutrition is a vital factor causing a high proportion of malaria-related mortality [12]. Also, malaria is strongly related to anaemia in children [13,14]. Though the relationship between malaria and malnutrition has shown some controversy, Sakwe *et al.* [15] found a significant interrelationship between malnutrition and malaria. But, due to malaria’s relationship with anaemia and malnutrition Teh *et al*. [16] recommended that reasonable control of anaemia and malnutrition will require adequate control of malaria infection. The anaemic child presents a more significant measure of undernutrition [15]. On the other hand, most importantly is the emergence of coexistence of these disease conditions and many other illnesses in an individual. These three health conditions cluster, but there is no know medical description for these interactions. Despite its complexity, the relationship between malaria, anaemia, and malnutrition is essential for our knowledge of childhood morbidity and the formulation of successful intervention methods [12].

In the past 20 years, Nigeria has not been able to meet the setting aside 15% of her national budget for health sector. The 2022 budget for health was less than 5% of the nation’s budget. This can hardly be enough to meet up with the realisations of the current national health policies [17]. The country’s low commitment to the socio-economic development that is the bane of the Sustainable Development Goals’ (SDGs) achievement means Nigeria is very unlikely to meet the SDGs by 2030. For instance, by 2019 index report, the country was ranked 159^th^ out of 162 countries in achieving SDGs so far [17]. This is even more worrisome now that the public health sector is being saddled with an emerging multimorbidity of childhood diseases. The identification and sharing of best practises to reduce health inequalities will be made possible by tracking global and national progress on the social determinants of health (SDH)[18]. In the light of the above, the principal contribution of the results from this study will help the policymakers to come up with informed decisions that will maximize the use of scarce resources by addressing multiple and cooccurrence of childhood diseases rather than independent approach. Furthermore, the Nigeria’s strategic position as the most populous black nation on earth is a global public health concerns because it has some of the lowest health indices in Africa. Due to its issues with development and quickly expanding population, the nation lowers the socioeconomic indicators for the entire continent of Africa [19]. Therefore, the lack of proper understanding of the causes, predictors, and prevention of these emerging public health conditions (multimorbidity) in children is of great concern particularly in the realization of the sustainable development goal-3 of ensuring healthy lives and promotion of well-being for all and especially in the associated aim of ending preventable deaths of new-borns and children by 2030.

To change the trajectory of providing adequate treatment for children living with multimorbidity, more research with a developing-country in focus is urgently needed to understand the causes and risk factors associated with multimorbidity in children in Nigeria. This study is a step in the right direction in unravelling the epidemiology and determinants of multimorbidity of common childhood diseases that will help government and policy makers and implementors to come up with an integrate model suitable for managing the multiple occurrences of diseases in children. Therefore, the purpose of this study was to investigate the multiple overlaps in the determinants of multimorbidity of anaemia, malaria, and malnutrition (MAMM) among children aged 6-59 months in Nigeria.

The specific objectives of this study include:

i. To describe the prevalence associated with some selected predictors, and spatial distributions of multimorbidity of anaemia, malaria, and malnutrition among children 6-59 months across Nigeria’s states and geopolitical regions using data from the 2018 NDHS.
ii. To investigate the individual and contextual risk factors of multimorbidity of malaria, anaemia, and malnutrition among children 6-59 months in Nigeria using data from the 2018 NDHS (with the incorporated contextual data from the National Human Development Report (2018 NHDR)).
iii. To determine the interaction effects of a child’s age, sex, and household socioeconomic status on the individual and contextual risk factors of MAMM among children aged 6-59 months in Nigeria.

## Materials and methods

### Variable Descriptions

#### The Dependent Variables

Three response (dependent) variables are considered in this study as indicators for a common childhood multimorbidity cluster: anaemia, malaria, and malnutrition statuses. These health conditions were captured objectively at the individual level in accordance with WHO recommended procedures, and the 2018 NDHS was the first time they have been captured simultaneously in any nationally representative health survey. The classifications of anaemia and malaria have been described elsewhere [20,21]. However, a measure of the overall description of malnutrition status among children aged 6-59 months in Nigeria was taken using the four indicators (stunting, wasting, underweight, and overweight). Children with no trace of anthropometric failure were classified as '0', labelled as 'well nourished', and those that have at least one of the four indicators were classified as '1', labelled as 'poorly nourished' [22–24].

#### The classification of multimorbidity Status

To classify multimorbidity across anaemia, malaria, and malnutrition among children aged 6- 59 months in the study sample, the 'Composite Index of Multi-morbidity' (CIMM), a technique adapted from the Composite Index of Anthropometric Failure (CIAF), was used [23,24]. The multimorbidity (with three outcomes) was classified into eight independent groups, such that it can have multi-categorical responses represented in three intersecting sets of anaemia, malaria, and malnutrition (Figure 1).

**Figure1:**
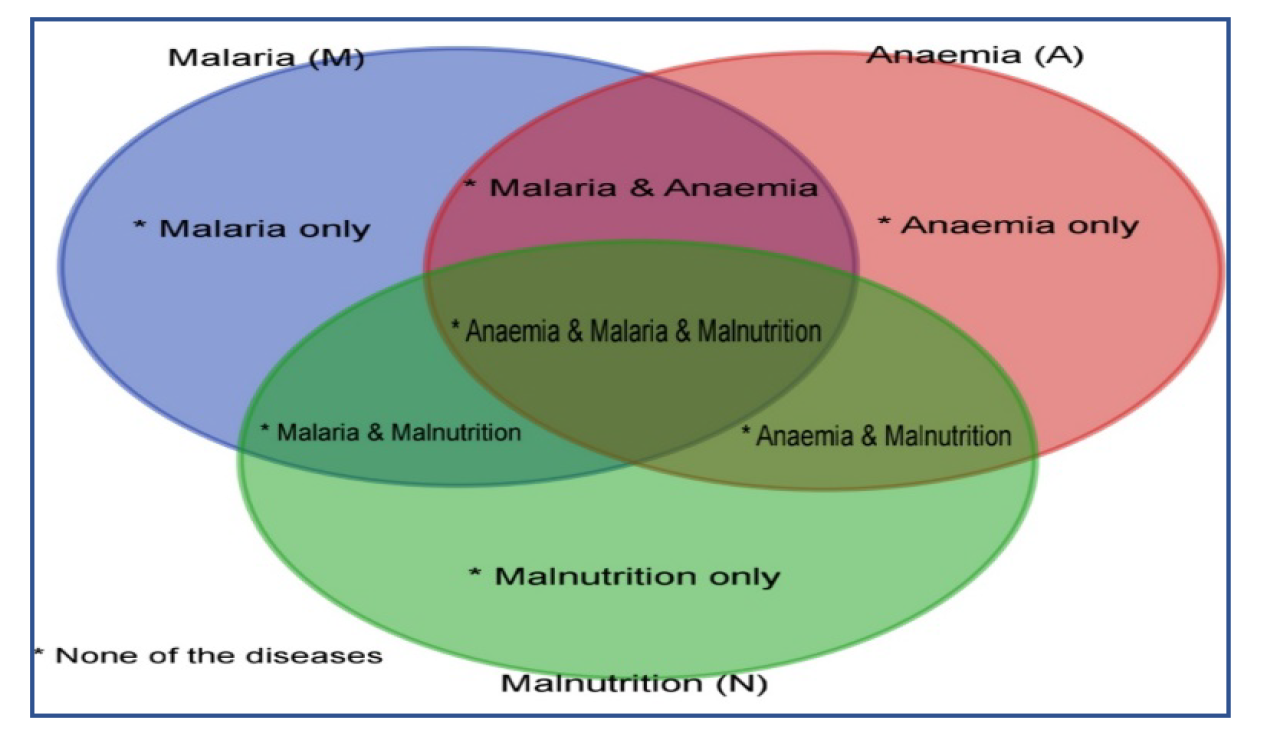
Diagram representing the intersection of the three outcome diseases

From the intersecting diseases (Figure 1), it recognises four distinct groups/categories: Those that had 'no disease' at all and are classified as '0'; those that had 'one disease only' and are classified as '1'; those that had two diseases only, and classified as '2', and those that had 'all three diseases, and classified as '3'. However, to align with the definition of multimorbidity as cooccurrence of two or more diseases in an individual, categories 2 and 3 were grouped into one (having two or more diseases). Therefore, the ordered set of multimorbidity conditions is S = {0, 1, 2}.

### Independent variables (Predictor variables)

The predictor variables considered for this study were categorised following earlier research and as presented in the NDHS 2018 final report in this study, the factors that affect a childhood's risk of contracting multimorbidity of anaemia, malaria, and malnutrition (MAMM) were broken down into factors related to the child, the parents, the households, and the area clusters.

### Analyses Techniques

#### Levels of statistical analyses

In the first level of analysis was the prevalence of MAMM across some variables and the spatial map descriptions across the states & FCT, and regions of residence in Nigeria. At the second level, a multilevel mixed effects ordered logistic regression models were fitted to determine the multiple overlaps in the variables that simultaneously predict the number of occurrences of MAMM among children 6-59 months of age in Nigeria. All the analyses were performed using Stata MP4 version 17 (StataCorp, College Station, USA), at 5% alpha level of statistical significance.

#### Ethical approval

The School of Health and Related Research (ScHARR) Ethics Committee of the University of Sheffield had given its permission for this research study's ethical conduct (Reference Number: 031534). Two nationally representative samples were used as the secondary analysis for this study. To use the data sets (2018 National Human Development Report and 2018 Nigeria Demographic and Health Survey), permission was sought from the Inner-City Fund (ICF)- International and the United Nations Development Program (UNDP-Nigeria), respectively. These two bodies (ICF-International and UNDP-Nigeria) have obtained formal consent to collect the data from the parent/guidance of the children.

## Results

### Prevalence of multimorbidity

The interactions between the three diseases (anaemia, malaria, and malnutrition) were considered as the state of multimorbidity of common childhood diseases clusters among children aged 6-59 months in Nigeria. Figure 2a shows that there were more children cohabiting with anaemia only, 22.5% (2293/10183), 95%CI (21.72-23.34), compared with malaria only, 3% (308/10183), 95%CI (2.71-3.38), and malnutrition only, 9% (897/10183), 95%CI (8.27-9.38). While in Figure 2b, the percentage of children with none of the outcome diseases, 'no disease' was 17.4% (1767/10183), 95%CI (16.63-18.10), while 48.3% (4917/10183), 95%CI (47.32-49.26) had two or more of the disease outcomes (multimorbidity), and 34.4% (3498/10183), 95%CI (33.44-35.29) had morbidity status (only one of the diseases)

**Figure 2:**
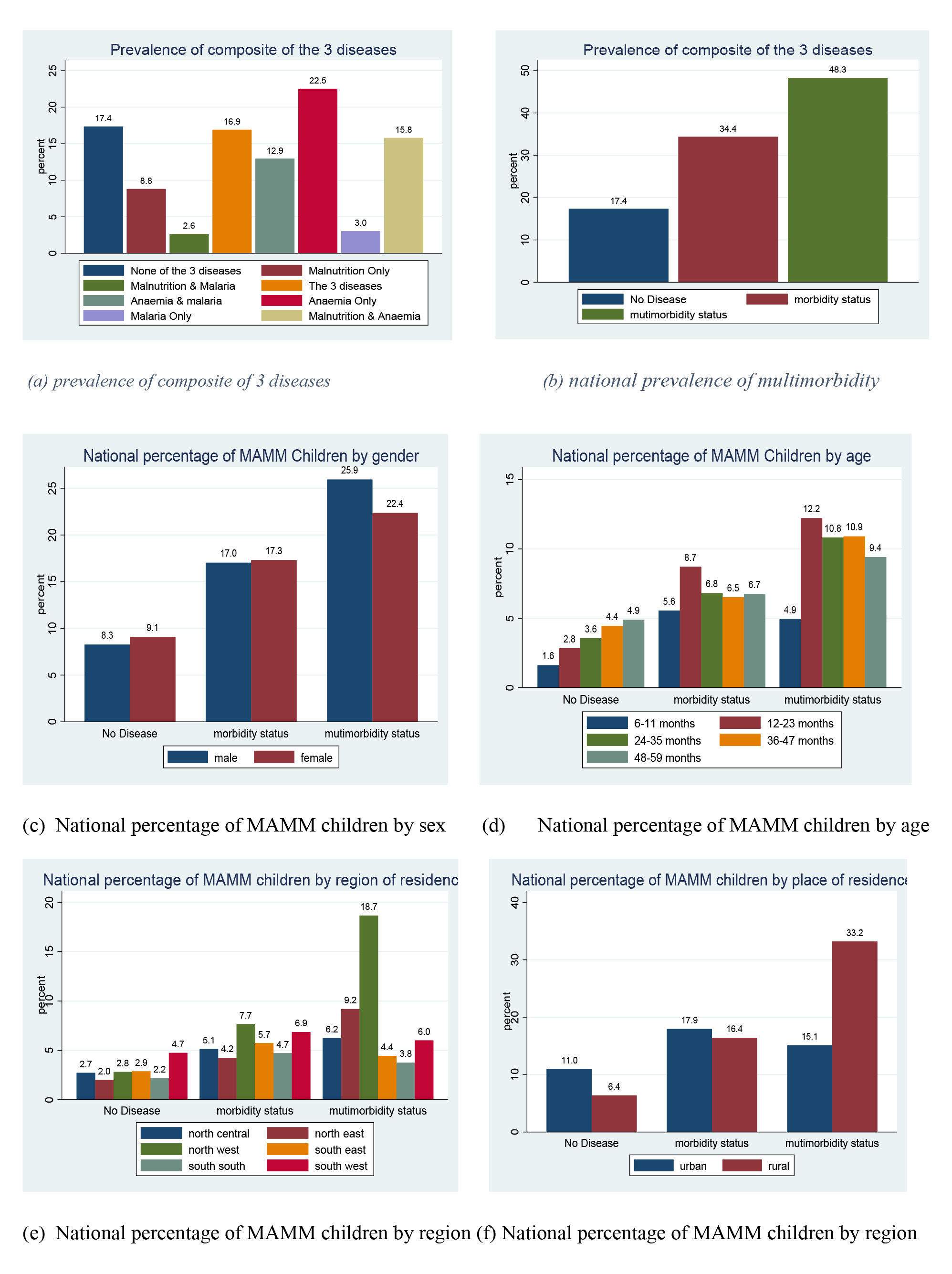
Distribution of National prevalence of MAMM across selected predictors Source: Data computed from Nigeria DHS 2018

**Figure 2:**
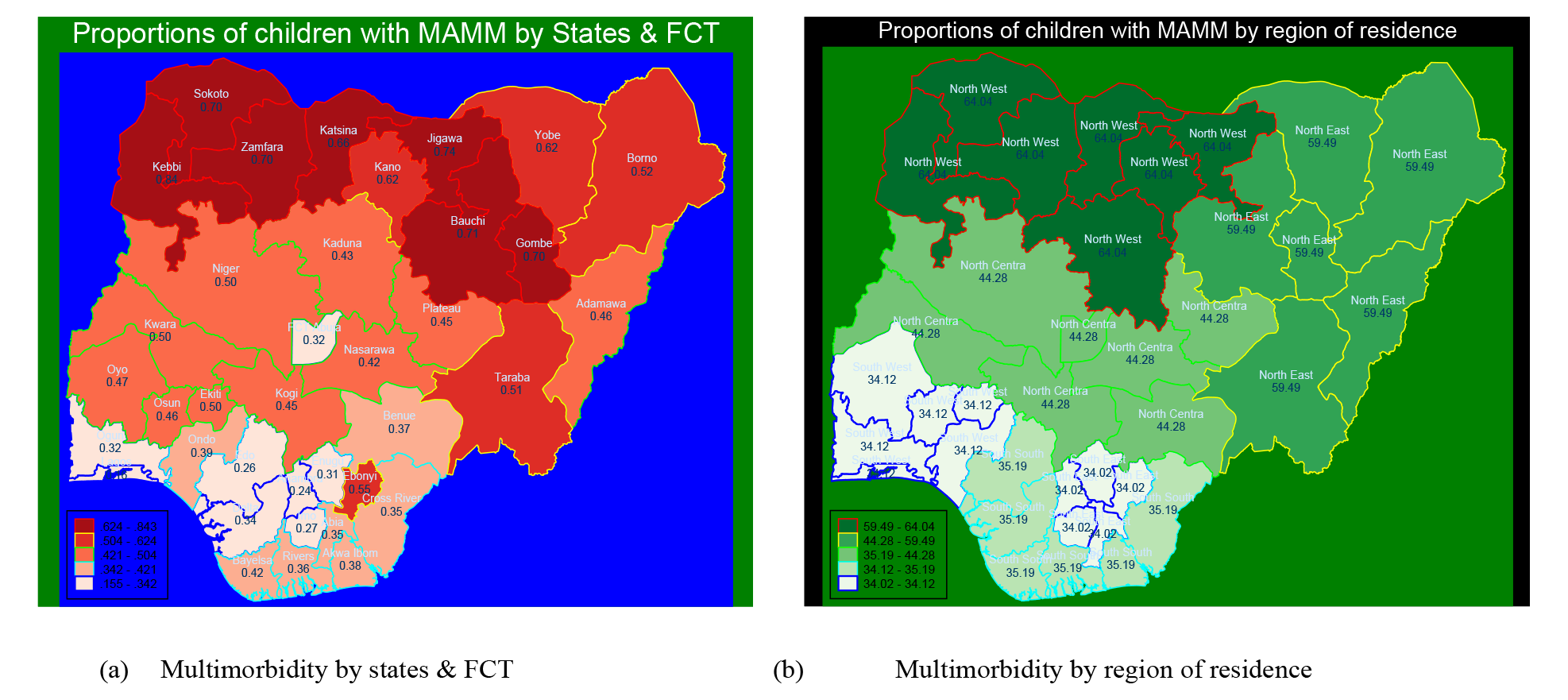
Spatial maps describing the proportions of children with two or more diseases Source: Data computed from Nigeria DHS 2018.

Figure 2c reveals that MAMM varies by sex. Male children were more prone to an increased MAMM prevalence than female children. Also, the prevalence of 'no disease' increases almost proportionally as age increases. The prevalence of MAMM was highest among children aged 12-23 months (Figure 2d). Similarly, in Figure 2e. the highest percentage of children aged 6- 59 months in Nigeria found sick of MAMM lived in the North-west geopolitical zones of Nigeria (18.7%), followed by children from the North-East geopolitical zone (9.2%). Also, in Fig 2f, the percentage of children in Nigeria cohabiting with two or more diseases of anaemia, malaria, and malnutrition from the rural area is more than two folds the number of those residing in an urban area.

#### Spatial proportions of the multimorbidity of two or more diseases by states and regions

Figure 3a presents the spatial variations in the proportion of multimorbidity of two or more diseases of anaemia, malaria, and malnutrition by states and FCT. It reveals that the proportion of children cohabiting with ‘2 or more diseases' in Nigeria was highest in Kebbi state with 0.83 (95% CI:0.78-0.86), followed by Jigawa state, 0.73 (95% CI: 0.69-0.78). Ebonyi state has the highest proportion, 0.55 (95% CI: 0.50-0.59), of children having concurrent two or more diseases of MAMM among the states in the southern part of Nigeria. Also, the map (Fig 3a) shows that the three states with the lowest proportions of MAMM in Nigeria are Edo state, 0.31 (95% CI: 0.24-0.40), Anambra state, 0.26 (95% CI: 0.22-0.31), and Lagos state, 0.14 (95% CI: 0.10-0.18). FCT has 0.36 (95% CI: 0.30-0.42).

**Figure 3:**
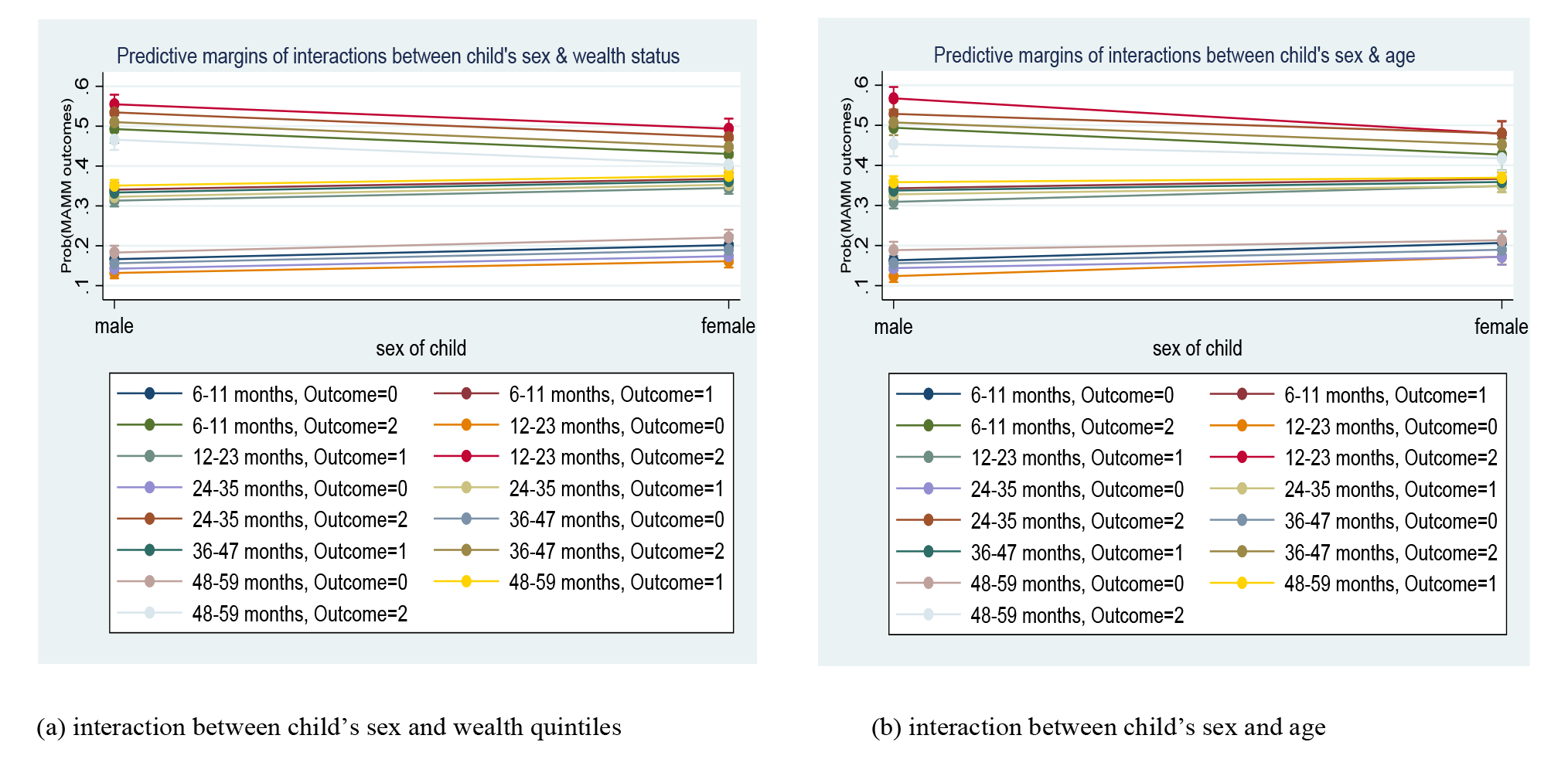
Predictive margins plot of interaction effects

Similarly, Figure 3b shows that children cohabiting with two or more diseases of anaemia, malaria, and malnutrition in North-west was the highest, 0.65 (95% CI: 0.62-0.67), followed by North-east geopolitical zones with a 0.59 (95% CI: 0.56-0.62) proportion of children living with MAMM in Nigeria. All the geopolitical zones in the southern part of Nigeria had similar distributions of multimorbidity and were below the national average of 0.48 (95% CI: 0.47- 0.49), including North-central with a proportion of 0.44 (95% CI: 0.41-0.47).

#### Multilevel analysis of multimorbidity status

A multicollinearity check using 'collin' command in STATA was done for all the predictor variables considered and 48 variables were extracted. Furthermore, three variables which formed the proxies for multimorbidity, anaemia, malaria, and malnutrition, were excluded before the next stage of the analysis was evaluated using variable selection methods to avoid over-or under-fitting across child-, parental-, and household-—community-, and area-related factors. This study recognises the shortfalls in using stepwise regression as a means of variable selection, but it remains the most popular method in the circle of researcher to date [25]. However, using these methods here is not an end to the analysis but a means of obtaining a model that will be most beneficial and economical [26]. To this end, applying the combine of backward and forward stepwise, 28 variables classified into child-related (child's sex, child's age in group, child's birth size, child's preceding birth interval, child took iron supplements, child's duration of breastfeeding, child took deworming drug in last 6months, child had fever in last 2 weeks before the survey, child's place of delivery), parental-related (maternal/caregiver highest educational level, mother is currently residing with husband/partner, mother's religious status, mother's anaemia status, maternal body weight status, paternal work status, partner education status), household-related (household wealth index, children under 5 slept under mosquito bed net last night, the sex of household head, number of people in household), community-related (proportion of community wealth level, proportion of community distance to health facility is no big problem, proportion maternal community education level), and area-related (multidimensional poverty index by state (MPI), human development index by state (HDI), gender inequality index by state (GII), geopolitical region of residence, and type of place of residence), were included in the analysis. The definition and classification of these variables have been reported elsewhere [20]

#### Multilevel mixed effect ordinal logistic regression models

The proportionality assumption was checked through a naïve method, showing that it was not violated. The predicted mean multimorbidity of the full model was 0.170, while the predicted mean multimorbidity of the partial model (after the variables that violated the proportionality assumptions were removed) was 0.166. The null hypothesis that the difference between these two means was not significantly different from zero at p<0.05 was not rejected (p=0.0729). So, it concludes that the proportionality assumption was not violated. Therefore, multilevel mixed effect ordinal logistic regression was used to answer research objective two.

In the first instance it was confirmed that the 2-level model was significantly nested within a 3-level model using a likelihood ratio test with χ^2^= 293.65, p<0.0001. Therefore, 3-level multilevel mixed effect ordinal logistic regression was carried out to evaluate this research objective.

## Model building

In this section, 5 models were evaluated. A weighted sample size of 7,794 children at level 1 nested in 1,360 communities at level 2, with an average of 6 children per community, and are, in turn, nested in 37 states at level 3 with an average of 210 children per state.

Table 1 shows that model 0 contained no covariate (variance component model), while models 1, 2, 3, and 4 considered multimorbidity of children aged 6-59 months in Nigeria as a function of child-related variables only, levels 1, 1&2, and 1, 2 & 3 predictors only, respectively. However, model 4, which contained all the level 1, 2, and 3 variables (full model), combined was adjudged the model of best fit with the highest log-likelihood (−6935.4) and least AIC (14010.9), therefore chosen for further analysis. The likelihood ratio test (1076.4), p<0.0001 of the variance component model indicates that the multilevel mixed effect ordinal logistic model is better than a single-level ordinal logistic model.

**Table 1:** Model fit comparison

| Model | No of covariates | Log-likelihood | AIC | BIC |
| --- | --- | --- | --- | --- |
| Model 0 | No covariate | -7415.6 | 14839.3 | 14867.1 |
| Model 1 | 9 (child-related) | -7254.5 | 14555 | 14715.1 |
| Model 2 | 20 (Level-1 factors only) | -6964 | 14026.1 | 14367.2 |
| Model 3 | 23 (level-1 & 2 factors only) | -6955.6 | 14015.4 | 14377.3 |
| Model 4 | 28 (level 1, 2, & 3 factors) | -6935.4 | 14010.9 | 14498.2 |

### Variance component analysis

Table 2 describes the variations in the prevalence of MAMM corresponding to the state (level 3), and community (level 2). For model 0 (null model), with no covariates, the variance component coefficient (VPC) at the state level is 0.099. meaning that differences across the state explain 10% of the total variability in the proportion of MAMM among children aged 6- 59 months (level 3). This is the same as the intrastate correlation coefficient (ICC), which signifies that the correlation between two children within the same state but in different communities is 0.099 (95% CI: 0.06-0.15).

**Table 2:**
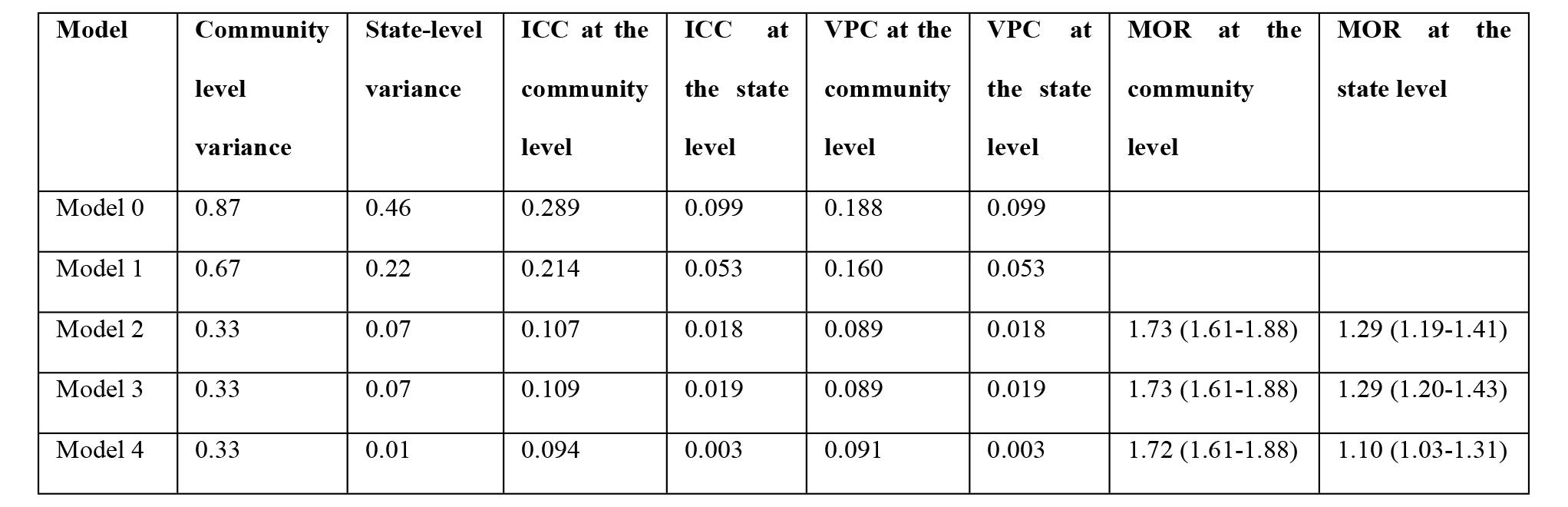
Distribution of random effect components

On the other hand, the VPC and ICC of model 0 at the community level (level 2), in a three-level model, are not usually the same (Obasohan et al 2021b). The VPC at level 2 (community) is 0.188, signifying that 18.8% of the total variability in the proportion of MAMM is attributable to the community. However, ICC at the community level is 0.289 (95% CI: 0.25- 0.33), suggesting that the correlation between two children/individuals (unit of analysis) within the same community and the same state is 28.9%. The sizes of ICCs at level-2 and level-3 are substantial (greater than 5%), which is large enough to validate further the need for a multilevel analysis over a single-level model [27].

Furthermore, from the chosen model (model 4), the intrastate and intracommunity correlations drop from 0.46 and 0.87 in the null model (model 0) to 0.01 and 0.33 in the model containing all 28 covariates, respectively. Also, between-state and between-community, the variabilities drop from 0.46 and 0.10 in the null model to 0.01 and 0.003 in the full-level model (containing level 1, 2, and 3 covariates), respectively. Again, this indicates that the distribution of all the variables across the states and communities differs significantly.

Additionally, a measure of odds for cluster variance was computed using the median odds ratios (Table 2). For the choice model (model 4), the median odds ratio (MOR) computed for states was 1.10, 95%CI (1.03-1.31) signifies a 10% increased risk of a child contracting '2 or more diseases' if he/she moves from one state to another with an increased risk of multimorbidity. Similarly, there is a 72% increased risk of a child contracting ‘2 or more diseases' if he/she moves to another community with a higher risk of MAMM.

## Fixed Effect Analysis

Table 3 reports the chosen model 4 (level 1, 2, & 3 covariates), and it reveals that the child's age, sex, birth size, pre-birth interval, the child was dewormed in the last six months before the survey, the child had a fever in the last two weeks before the survey, the maternal education status, religious status, anaemia status, body weight index status, paternal education status, the household wealth status, median proportion of people in the community who said distance to a health facility is no big problem, state human development index (HDI), state gender inequality index (GII), region of residence, and place of residence were significant predictors of the proportional odds of a child cohabiting with MAMM versus the combined of 0/1 diseases among children 6-59 months of age in Nigeria compared with their respective reference category.

**Table 3:** Multilevel Ordinal logistic regression analysis of the individual, community, and state level risk factors for MAMM

|  | Model 2 (N=7794)<br>(Level 1 covariates only) |  | Model 3 (N=7794)<br>(Level 1 & 2 covariates only) |  | Model 4 (N=7794)<br>(Level 1, 2, & 3 covariates) |  |
| --- | --- | --- | --- | --- | --- | --- |
| Variables | AOR | 95% CI | AOR | 95% CI | AOR | 95% CI |
| **Child's sex** |  |  |  |  |  |  |
| Male | 1.00 |  | 1.00 |  | 1.00 |  |
| Female | 0.73*** | (0.67-0.8) | 0.73*** | (0.67-0.81) | 0.73*** | (0.67-0.80) |
| **Child's age in group** |  |  |  |  |  |  |
| 6-11 months | 1.00 |  | 1.00 |  | 1.00 |  |
| 12-23 months | 1.39*** | (1.18-1.65) | 1.39*** | (1.18-1.65) | 1.39*** | (1.18-1.65) |
| 24-35 months | 1.26* | (1-1.57) | 1.24 | (0.99-1.56) | 1.25* | (1.00-1.57) |
| 36-47 months | 1.10 | (0.87-1.38) | 1.09 | (0.87-1.37) | 1.1 | (0.88-1.38) |
| 48-59 months | 0.88 | (0.7-1.1) | 0.87 | (0.69-1.09) | 0.87 | (0.69-1.10) |
| **Child's birth size** |  |  |  |  |  |  |
| Large | 1.00 |  | 1.00 |  | 1.00 |  |
| Average | 0.99 | (0.84-1.16) | 0.99 | (0.84-1.16) | 0.98 | (0.83-1.15) |
| Small | 1.26* | (1.02-1.56) | 1.26* | (1.02-1.56) | 1.26* | (1.02-1.56) |
| **Preceding birth interval** |  |  |  |  |  |  |
| None | 1.00 |  | 1.00 |  | 1.00 |  |
| 8-24 months | 1.36*** | (1.16-1.6) | 1.37*** | (1.17-1.61) | 1.37*** | (1.17-1.62) |
| 25-35 months | 1.09 | (0.94-1.28) | 1.10 | (0.94-1.29) | 1.11 | (0.95-1.29) |
| 36-59 months | 1.01 | (0.86-1.18) | 1.01 | (0.86-1.19) | 1.02 | (0.87-1.20) |
| 60+ months | 0.93 | (0.75-1.14) | 0.93 | (0.76-1.14) | 0.93 | (0.76-1.15) |
| **Child took Iron supplements** |  |  |  |  |  |  |
| No | 1.00 |  | 1.00 |  | 1.00 |  |
| Yes | 1.04 | (0.91-1.19) | 1.04 | (0.91-1.19) | 1.03 | (0.90-1.18) |
| **Duration of breastfeeding** |  |  |  |  |  |  |
| Ever breastfed, not currently breastfeeding | 1.00 |  | 1.00 |  | 1.00 |  |
| Never breastfed | 1.23 | (0.83-1.82) | 1.20 | (0.81-1.79) | 1.19 | (0.80-1.76) |
| Still breastfeeding | 0.96 | (0.81-1.15) | 0.95 | (0.8-1.14) | 0.96 | (0.80-1.14) |
| **Child was dewormed in last 6 months before the survey** |  |  |  |  |  |  |
| No | 1.00 |  | 1.00 |  | 1.00 |  |
| Yes | 0.83** | (0.73-0.94) | 0.83** | (0.73-0.94) | 0.84** | (0.74-0.96) |
| **Child had fever in last 2 weeks before the survey** |  |  |  |  |  |  |
| No |  |  |  |  | 1.00 |  |
| Yes | 1.56*** | (1.39-1.75) | 1.56*** | (1.39-1.75) | 1.57*** | (1.40-1.77) |
| **Place of child's delivery** |  |  |  |  |  |  |
| Home | 1.00 |  | 1.00 |  | 1.00 |  |
| Public facility | 0.90 | (0.79-1.03) | 0.92 | (0.8-1.05) | 0.93 | (0.81-1.06) |
| Private facility | 0.88 | (0.74-1.04) | 0.89 | (0.75-1.05) | 0.88 | (0.74-1.04) |
| Elsewhere | 1.03 | (0.72-1.46) | 1.02 | (0.72-1.46) | 1.01 | (0.71-1.44) |
| **Maternal/caregiver highest educational level** |  |  |  |  |  |  |
| No education |  |  |  |  | 1.00 |  |
| Primary | 0.83** | (0.7-0.99) | 0.84* | (0.7-1) | 0.84 | (0.70-0.99) |
| Secondary | 0.69*** | (0.58-0.82) | 0.69*** | (0.57-0.83) | 0.68*** | (0.57-0.82) |
| Higher | 0.45*** | (0.35-0.57) | 0.45*** | (0.34-0.58) | 0.45*** | (0.35-0.58) |
| **Mother staying with a partner** |  |  |  |  |  |  |
| Staying with partner | 1.00 |  | 1.00 |  | 1.00 |  |
| staying elsewhere | 0.94 | (0.77-1.14) | 0.94 | (0.78-1.15) | 0.94 | (0.77-1.14) |
| **Mother's religious status** |  |  |  |  |  |  |
| Catholic | 1.00 |  | 1.00 |  | 1.00 |  |
| Other Christian | 1.08 | (0.9-1.3) | 1.10 | (0.91-1.32) | 1.12 | (0.93-1.36) |
| Islam | 1.23 | (0.99-1.53) | 1.28* | (1.02-1.6) | 1.37** | (1.08-1.74) |
| Traditionalist & others | 0.96 | (0.54-1.71) | 0.97 | (0.54-1.71) | 1 | (0.56-1.78) |
| **Mother's Anaemia status** |  |  |  |  |  |  |
| Not Anaemic |  |  |  |  | 1.00 |  |
| Anaemic | 1.61*** | (1.46-1.78) | 1.61*** | (1.46-1.78) | 1.6*** | (1.45-1.77) |
| **Maternal body mass index** |  |  |  |  |  |  |
| Normal | 1.00 |  | 1.00 |  | 1.00 |  |
| Underweight | 1.16 | (0.97-1.38) | 1.16 | (0.98-1.39) | 1.18 | (0.99-1.40) |
| Overweight | 0.76*** | (0.67-0.86) | 0.76*** | (0.67-0.87) | 0.77*** | (0.67-0.87) |
| Obese | 0.70*** | (0.59-0.83) | 0.71*** | (0.6-0.84) | 0.71*** | (0.60-0.84) |
| **Paternal work status** |  |  |  |  |  |  |
| Not working | 1.00 |  | 1.00 |  | 1.00 |  |
| Working | 1.24 | (0.94-1.66) | 1.23 | (0.92-1.64) | 1.21 | (0.91-1.61) |
| **Partner education status** |  |  |  |  |  |  |
| No education | 1.00 |  | 1.00 |  | 1.00 |  |
| Primary education | 0.97 | (0.81-1.17) | 0.99 | (0.82-1.19) | 0.97 | (0.81-1.17) |
| Secondary education | 0.88 | (0.74-1.05) | 0.90 | (0.76-1.06) | 0.9 | (0.76-1.07) |
| Tertiary education | 0.79** | (0.64-0.98) | 0.81 | (0.65-1) | 0.81* | (0.66-1.00) |
| **Household wealth index** |  |  |  |  |  |  |
| Poorest | 1.00 |  | 1.00 |  | 1.00 |  |
| Poorer | 0.82** | (0.69-0.99) | 0.86 | (0.72-1.04) | 0.86 | (0.72-1.04) |
| Middle | 0.58*** | (0.48-0.71) | 0.69*** | (0.56-0.85) | 0.69*** | (0.56-0.86) |
| Richer | 0.43*** | (0.35-0.53) | 0.54*** | (0.43-0.69) | 0.56*** | (0.44-0.72) |
| Richest | 0.28*** | (0.22-0.36) | 0.36*** | (0.27-0.47) | 0.38*** | (0.29-0.50) |
| **Under-5 slept under a bed net** |  |  |  |  |  |  |
| No child | 1.00 |  | 1.00 |  | 1.00 |  |
| All children | 0.94 | (0.8-1.1) | 0.93 | (0.79-1.09) | 0.93 | (0.79-1.09) |
| Some children | 1.12 | (0.91-1.38) | 1.11 | (0.9-1.37) | 1.12 | (0.91-1.39) |
| No net in household | 0.92 | (0.79-1.09) | 0.93 | (0.79-1.09) | 0.94 | (0.80-1.11) |
| **Sex of household head** |  |  |  |  |  |  |
| Male | 1.00 |  |  |  | 1.00 |  |
| Female | 0.96 | (0.79-1.18) | 0.95 | (0.78-1.17) | 0.95 | (0.78-1.16) |
| **Number of people in household** |  |  |  |  |  |  |
| 2-3 | 1.00 |  | 1.00 |  | 1.00 |  |
| 4-6 | 0.89 | (0.73-1.08) | 0.89 | (0.73-1.07) | 0.88 | (0.73-1.07) |
| 7-9 | 0.99 | (0.8-1.22) | 0.98 | (0.79-1.22) | 0.97 | (0.78-1.20) |
| 10+ | 1.14 | (0.9-1.44) | 1.13 | (0.89-1.43) | 1.12 | (0.88-1.41) |
| **Median community wealth level** |  |  |  |  |  |  |
| Low |  |  | 1.00 |  | 1.00 |  |
| High |  |  | 0.74** | (0.61-0.9) | 0.79* | (0.65-0.96) |
| **Median proportion of community distance to health facility is no big problem** |  |  |  |  |  |  |
| Low |  |  | 1.00 |  | 1.00 |  |
| High |  |  | 0.86* | (0.75-0.98) | 0.86* | (0.75-0.98) |
| **Media proportion of community maternal education level** |  |  |  |  |  |  |
| Low |  |  | 1.00 |  | 1.00 |  |
| High |  |  | 1.10 | (0.91-1.33) | 1.10 | (0.90-1.34) |
| **State multidimensional poverty index (SMPI)** |  |  |  |  |  |  |
| Highly deprived |  |  |  |  | 1.00 |  |
| Above averagely deprived |  |  |  |  | 1.28 | (0.95-1.73) |
| Averagely Deprived |  |  |  |  | 0.72 | (0.48-1.07) |
| Mildly Deprived |  |  |  |  | 0.69 | (0.45-1.06) |
| Lowest Deprived |  |  |  |  | 0.67 | (0.41-1.11) |
| **State human development index (HDI)** |  |  |  |  |  |  |
| Lowest HDI |  |  |  |  | 1.00 |  |
| Low HDI |  |  |  |  | 1.25 | (0.94-1.65) |
| Average HDI |  |  |  |  | 1.27 | (0.90-1.80) |
| High HDI |  |  |  |  | 1.51* | (1.01-2.28) |
| Highest HDI |  |  |  |  | 1.13 | (0.72-1.78) |
| **Gender inequality index by state (GII)** |  |  |  |  |  |  |
| Lowest GII |  |  |  |  | 1.00 |  |
| Low GII |  |  |  |  | 0.76* | (0.58-1.00) |
| Average GII |  |  |  |  | 1.25 | (0.94-1.67) |
| High GII |  |  |  |  | 1.03 | (0.80-1.32) |
| Highest GII |  |  |  |  | 1.30 | (0.94-1.80) |
| **Region of residence** |  |  |  |  |  |  |
| North-central |  |  |  |  | 1.00 |  |
| North-east |  |  |  |  | 0.68* | (0.49-0.96) |
| North-west |  |  |  |  | 1.12 | (0.77-1.62) |
| South-east |  |  |  |  | 1.56** | (1.13-2.15) |
| South-south |  |  |  |  | 1.52** | (1.09-2.12) |
| South-west |  |  |  |  | 1.61*** | (1.16-2.23) |
| **Type of place of residence** |  |  |  |  |  |  |
| Urban |  |  |  |  | 1.00 |  |
| Rural |  |  |  |  | 1.23*** | (1.06-1.42) |

*AOR: Adjusted odds ratios, CI: Confidence interval, \* p < 0.05, \*\* p < 0.01, \*\*\* p < 0.001*

### Individual-level predictors of multimorbidity

From model 4 in Table 3, the proportional odds of female children cohabiting with '2 or more diseases' versus the combined of '0/1 disease only' is 0.27 (95% CI: 0.67-0.80, p=0.001), lower when compared to male children, relative to other predictors being held constant. Likewise, the proportional odds of children aged 12-23 months having 'two or more diseases' versus a combined of '0/1 diseases' is 1.39 (95% CI: 1.18-1,65), times higher when compared to children aged 6-11 months with all other variables held constant. A child born as perceived small size by the mother had statistically significant proportional odds of 1.26 (95% CI: 1.02-1.56, p=0.04), times of having at least 'one disease' versus 'no disease' compared to children who were born perceived large by their mothers while accounting for other predictors.

Furthermore, children whose mothers have higher education levels have a more than two folds reduced proportional odds (AOR=0.45, 95% CI: 0.35-0.58) of cohabiting with '2 or more diseases' versus combined odds of having '0/1 disease' when compared with children whose mothers do not have any formal education. Also, children of Muslim mothers had significant proportional odds of 0.37 (95% CI: 1.08-1.74), more of contracting '2 or more diseases' versus '0/1 disease' compared with children whose mothers are Catholics. However, the proportional odds were not significant in a model with only level-1 covariates (model 2). Additionally, children whose mothers are anaemic; overweight, or obese had 0.6 increased odds, 0.23, or 0.29 reduced odds, of cohabiting with '2 or more diseases' versus a combined odds of '0/1 disease', respectively, when compared with children whose mothers are neither 'not anaemic' nor have 'normal body mass index'.

For household wealth quintiles, the results reveal that the higher the wealth quintile, the less likely the proportional odds of the children 6-59 months of age from the households of contracting '2 or more diseases' versus the combined odds of '0/1 disease'. Children from the middle, richer, and richest classes have reduced statistically significantly proportional odds of 0.31, 0.44, and 0.62, respectively, of being sick of '2 or more diseases' versus combined odds of '0/1 disease' when compared with children from the poorest household wealth quintile.

Out of the three community (level 2) covariates in this analysis, two were statistically significant predictors of MAMM among children 6-59 months of age in Nigeria. Children from a community where the proportion of the wealth level is high (median and above) had reduced proportional odds of 0.79 (95% CI: 0.65-0.96), times of contracting multimorbidity of '2 or more diseases' versus 0/1 disease when compared with children from a community with low (less than the median) wealth level. Similarly, children living in a community where the median proportion of the people where community distance to a health facility is 'no big problem' is high (median and above) have 0.14 reduced proportional odds of contracting '2 or more diseases' versus '0/1 disease' compared with other children from the communities with low proportion relative to other covariates in the model.

Moreover, out of the five state-level covariates adjusted for in model 4 analysis, state multidimensional poverty index (SMPI) was not a statistically significant predictor of MAMM among children 6-59 months of age in Nigeria, conditional other covariates. However, children from the states where the human development index is in the high quintile have statistically significant harmful proportional odds of contracting '2 or more diseases' versus 0/1 disease when compared with children from the state where human development index is at the lowest quintile. Similarly, children from the states where the gender inequality index (GII) is low have reduced proportional odds of 0.24 of contracting multimorbidity versus '0/1 disease' when compared with children from the state where GII is the lowest quintile. Also, concerning the region of residence, only the proportional odds of contracting '2 or more diseases' versus the combined of '0/1 disease' for children from the North-west is not statistically significant when compared with children from the North-central geopolitical zone of Nigeria. Finally, children from rural areas have increased proportional odds of 0.23 (95% CI: 1.06-1.42), more of cohabiting with '2 or more diseases' versus the combined odds of '0/1 disease' when compared with children from urban areas.

## Interaction effects

Additionally, the interaction effects of a child's age, sex, and household socioeconomic status on the individual and contextual risk factors of MAMM among children aged 6-59 months in Nigeria, were further investigated conditional on the model 4 (full model) covariates. In a scoping review of multimorbidity of childhood diseases, it was found that child's age, sex, and household wealth status were the most common predictors of all the diseases examined in the extracted papers [28]. The three individual-level variables were classified into four possible interaction groups to include three, ‘2-way' and one, ‘3-way' classifications: child's sex and wealth status; child's sex and age; child's age and wealth status; and child's sex, age, and wealth status. In view of this, five additional models (models 5– 9) were created. Model 5 includes model 4 conditional the interaction between child's sex and wealth status; model 6 includes model 4 and interaction of child's sex and age; model 7 involves model 4 and the interactions between child's age and wealth status; model 8 has model 4 while accounting the three 2-way interactions of child's sex, age, and household wealth status. Finally, model 9 includes model 8 and the 3-way interactions between a child's sex, age, and household wealth quintiles. Two approaches could be used simultaneously to interpret the interaction effects. (i) By examining the significance of the interaction terms (for detailed results see S1 Appendix) (ii) By visual inspection of the interaction plots. Here, the interpretations were provided using the plots. Indeed, interaction plots are necessary for a more straightforward interpretation [29]. The parallel lines in Figure 3a and 3b indicate the non-presence of interaction effects between a child's sex and household wealth quintiles, and between child's age and sex, respectively. The implications are that the effect of a child's sex on MAMM does not vary by the household wealth quintile as well as child's age. In other words, the household wealth status and child's age do not moderate the relationship between the child's sex and MAMM.

Sometimes, interpretations of interaction effects could be challenging for odds ratios in ordinal logistic regression models [30]. However, an intuitive way to present this seems complicated to understand the situation [31,32] is here presented in Table 4 and Figure 4 showing the output of margins of the predictive effect of interaction between child's age and wealth status and MAMM after adjusting for model 4 covariates. Figure 4 shows that some of the lines' intersections (non-parallel) indicate the existence of interaction effects. For instance, in the band of 'none of the diseases' (bottom) and '2 or more diseases' (top), the effects of the richest household vary at different points in the child's age band. Table 4 and Figure 4, in the 'none of the diseases' (healthy) group, a child living in the richest household and aged 48-59 months has a probability of 0.31 of staying healthy of MAMM compared with a child of the same age but resides in the poorest household with a probability 0.12 of being healthy from MAMM.

**Figure 4.**
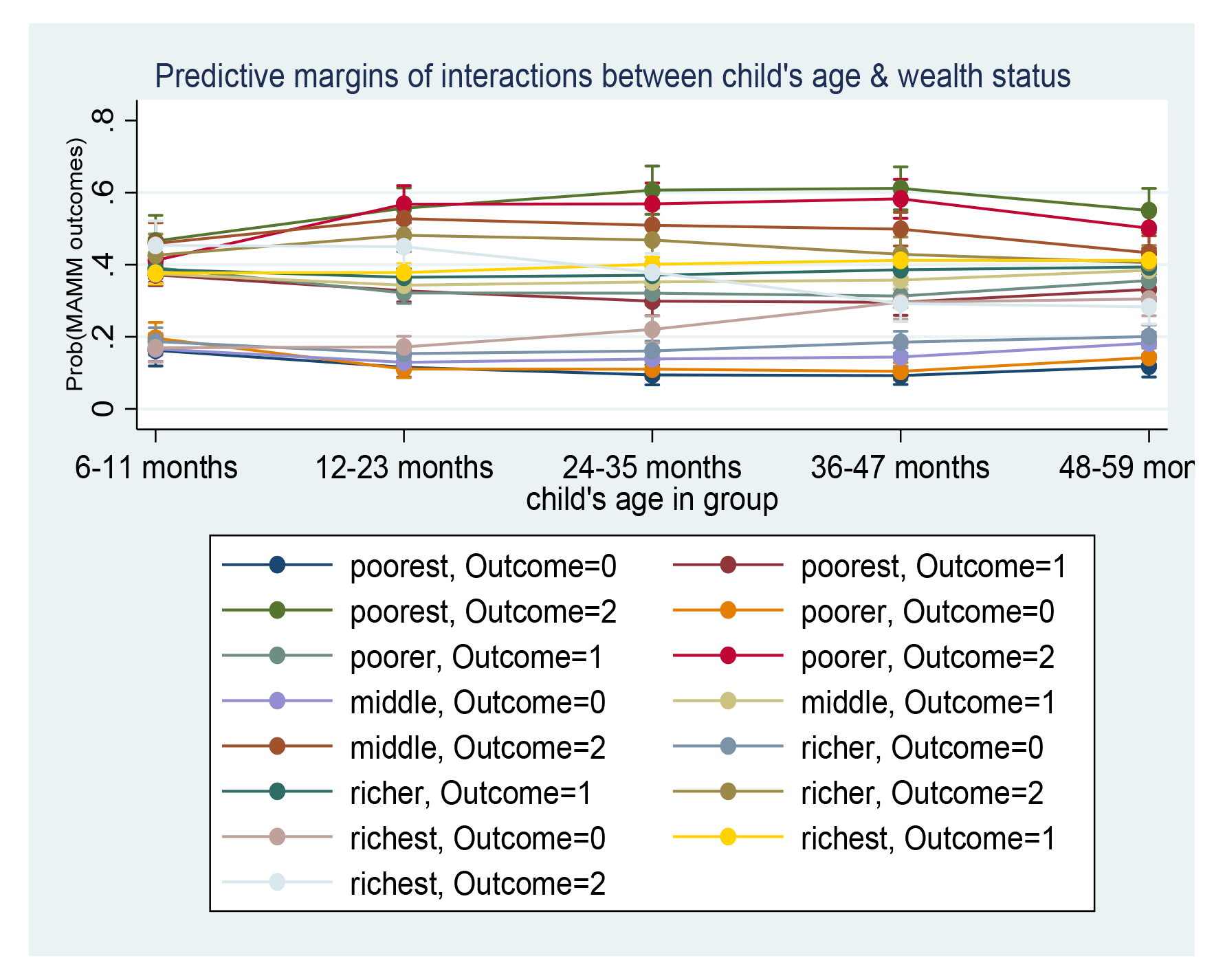
Predictive margins plot of interaction effects of child's age & wealth status

**Table 4.** Average adjusted probability for interactions of child's age and wealth status

|  | None of the disease |  |  |  |  | One disease only |  |  |  |  | Two or more diseases |  |  |  |  |
| --- | --- | --- | --- | --- | --- | --- | --- | --- | --- | --- | --- | --- | --- | --- | --- |
|  | 06-11 | 11-23 | 24-35 | 36-47 | 48-59 | 06-11 | 11-23 | 24-35 | 36-47 | 48-59 | 06-11 | 11-23 | 24-35 | 36-47 | 48-59 |
| **Poorest** | 0.16 | 0.12 | 0.09 | 0.09 | 0.12 | 0.37 | 0.33 | 0.30 | 0.30 | 0.33 | 0.47 | 0.56 | 0.61 | 0.61 | 0.55 |
| **Poorer** | 0.20 | 0.11 | 0.11 | 0.10 | 0.14 | 0.39 | 0.32 | 0.32 | 0.31 | 0.36 | 0.41 | 0.57 | 0.57 | 0.58 | 0.50 |
| **Middle** | 0.17 | 0.13 | 0.14 | 0.14 | 0.18 | 0.38 | 0.34 | 0.35 | 0.36 | 0.39 | 0.46 | 0.53 | 0.51 | **0.50\*** | **0.43\*** |
| **Richer** | 0.19 | 0.15 | 0.16 | 0.19 | 0.20 | 0.39 | 0.37 | 0.37 | 0.39 | 0.39 | 0.43 | 0.48 | 0.47 | **0.43\*** | 0.41 |
| **Richest** | 0.17 | 0.17 | 0.22 | 0.30 | 0.31 | 0.38 | 0.38 | 0.40 | 0.41 | 0.41 | 0.45 | 0.45 | **0.38\*** | **0.29\*** | **0.28\*** |

Similarly, children aged 36-47 months living in the poorest households have the probability of contracting MAMM of 0.11, 0.18, and 0.32 above the probability of children of the same age who live in the middle, richer, and richest households while all other model 4 covariates are held constant. Furthermore, in the 'two or more diseases' (MAMM) group, a child living in the richest household and aged 48-59 months has a significantly lower probability of 0.28 of contracting MAMM compared with a child of the same age but resides in the poorest household with a probability 0.55 of cohabiting with MAMM.

## Discussion

This current study focuses on multimorbidity of the three most common objective childhood cluster diseases using anaemia, malaria, and malnutrition as proxies for disease cooccurrence. A three-state multilevel mixed effect ordinal logistic regression was used to find the significant predictors of cohabiting with two or more diseases among children aged 6-59 months in Nigeria. Multimorbidity studies in children had been neglected for a long time, especially in the LMICs. It is challenging to evaluate this study's findings in light of earlier research, partly because of the lack of similar studies, differences in methodological approach, the disease conditions, survey types and population settings [33]. Nevertheless, this study has found significant disparities in some child-, parental-, household-, community-, and area-related risk factors on the occurrence of MAMM.

This current study found approximately a two-fold prevalence of 48.3% of MAMM among children aged 6-59 months in Nigeria, compared with the highest prevalence in an earlier scoping review (Obasohan et al 2023). Also, there are more children in Nigeria cohabiting with two or more of MAMM than those cohabiting with one disease of anaemia, malaria, and malnutrition, or none of the diseases.

Similarly, spatial disparities in the proportion of children aged 6-59 months in Nigeria cohabiting with '2 or more diseases' were found across the regions and states in Nigeria. This was highest in Kebbi state, followed by Jigawa state. Also, three states, Edo state, Anambra, and Lagos states have estimated proportions of 0.31 (95% CI: 0.24-0.40), 0.26 (95% CI: 0.22-0.31), and 0.14 (95% CI: 0.10-0.18). respectively. Furthermore, North-west had the highest proportion of children with multimorbidity across the region of residence in Nigeria. All the geopolitical zones in the southern part of Nigeria (South-West, South-East, and South-South) have similar multimorbidity distributions, below the national average of 0.48 (95% CI: 0.47- 0.49).

The findings showed child's sex is significantly protective for female children in MAMM compared to their male counterparts while holding other predictors constant. This finding implies that female children are less likely to co-inhabit with MAMM than male children when other covariates are held constant. This finding is supported by previous studies, which revealed that female children are less likely to cohabit with multimorbidity of childhood diseases [34,35], but the finding is contrary in Tran et al. [36] who concluded that it was more likely for a female to cohabit with '2 or more diseases. Similarly, the older the children become, the more likely they live with MAMM. Children aged between the 1st and 3^rd^ year are at significantly increased risk of cohabiting with MAMM compared to children less than one year.

This finding is supported by a study conducted by Adedokun [37] in a similar setting. Also, [35,36] supported the result, but contrary results that being older is protective for children under-five years compared with children less than one year were reported [34,38,39]. The findings of this study also indicated that children born small-sized were more likely to cohabit with MAMM than large-sized children at birth. This finding did not support the conclusion made by [37], who reported that children born big are likely to have an increased risk of multimorbidity of fever, pneumonia, and diarrhoea. The contrary findings are because the two studies have used different disease conditions as a proxy for multimorbidity. This study also demonstrated that children aged 6-59 months in Nigeria who experienced a fever two weeks before the survey date were much more at risk of having two or more MAMM than children who did not. This result is in line with findings from other studies [34].

The findings also show that maternal education, maternal religion is Islam, maternal anaemia status, and body mass index were significantly related to MAMM. On maternal educational attainment, the results supported the hypothesis that maternal education has a negative correlation with children's health outcomes [37]. Moreover, children of mothers with higher educational status had a lower risk of multimorbidity—defined as having 'two or more of anaemia, malaria, and malnutrition compared with children of mothers with no formal education conditional upon holding other predictors constant. This finding is consistent with the conclusions of other research [35,36] but disagreed with the findings in [37,39]. In addition, children whose fathers had higher education exhibited protective effects from MAMM. More excellent education for the mothers, particularly the caregivers, often improved their knowledge, attitude, and practice of typical children's diseases [20].

Also, the finding shows that children whose mothers were anaemic are at an increased risk of cohabiting with two or more MAMM. The possible explanation from previous studies is that children of anaemic mothers are more likely to be anaemic and are at high risk of poor child,health outcomes [21,40]. This current study also found that children of mothers who are overweight or obese have decreased significant odds of contracting two or more MAMM versus 0/1 disease compared to children whose mothers have normal body mass index when other covariates are held constant. Also, mothers' underweight status harms their children (though not significant). Like their mothers, underweight children are more likely to be underweight and may experience adverse effects on their development and health [41]. The results of this study regarding the maternal weight spectrum contradict the theory that developing nations may be at increased risk for both underweight and overweight [41] and have shown that overweight mother status is protective while underweight mother status is harmful. Also, according to this study, maternal obesity protects the neonate, and the mothers in this group are more likely to come from wealthy households in developing nations [41]. To accurately account for the actual position, more study is needed.

Of the number of household-related variables considered in this study which includes household wealth quintiles, number of children under-five years that slept under a bed net the night before the survey, sex of household head, and the number of persons in a household, only household wealth appears significant predictor of MAMM when other predictors are held constant. A higher household socioeconomic level had a lower risk of developing '2 or more of MAMM. This finding agrees with the conclusions of [36,37,39]. However, children from lower-income families are more likely to contract childhood diseases. This result relates to the fact that low-income families may find it challenging to purchase nutritious food that will increase their intake of nutrients and help them develop immunity to diseases [37]. They may also live in impoverished or densely populated areas [42]] and may have more children than they can reasonably care for [43], which is likely to increase the risk of multimorbidity in children.

This current study found that the higher (median and above) the proportion of community wealth status, the more protective the children become of MAMM. This finding supports other studies that children from low-income households or communities have poor health outcomes [44]. Similarly, this study reports that the higher the proportion of those in the community who affirmed that the distance to the nearest health centre is ‘no big problem', the less likely the children from such a community will contract two or more of MAMM. Distance to health centres could be a significant factor in getting prompt medical attention, and delays in getting treatment for paediatric illnesses could have more severe consequences. Oldenburg et al. agree that children that have less access to primary care may be more susceptible to poor health outcomes, including mortality as a result, they may be a population for which initiatives aimed at reducing child mortality and morbidity should be prioritised [45]. Some studies disagree with this finding [46,47].

On the area-related predictors, five variables were considered: state multidimensional poverty index (MDPI), state human development index (HDI), state gender inequality index (GII), region of residence, and place of residence. Though MDPI was not a significant predictor of MAMM, the results were exciting and informative. Children living in the above-averagely deprived state are at a higher risk of MAMM compared with children in highly deprived. Conversely, children living in the state that are averagely, mildly, and lowest deprived had reduced odds of cohabiting with MAMM when all other covariates are held constant. The non-significance of MDPI in a child's health outcomes was supported by another study that found mixed conclusions in MDPI indicators at the state level [48]. Surprisingly, this current study found that children from states with high human development index were significantly more likely to cohabit with MAMM than those living in states with the lowest HDI. This finding is contrary to the general expectation that the higher the state HDI, the less likely the children from the such a state will cohabit with adverse health outcomes [49]. The possible explanation for this result is that 14 out of 37 states were classified as 'high HDI', leading to high variability in MAMM in this group, compared with six states classified as 'lowest HDI' with low variability in MAMM. The high heterogeneity in MAMM across these 14 states must have resulted in their weak effects and significance. So, a non-linear impact of HDI on a child's health outcomes is therefore suspected [50]. The reasons for this happening can be explored further in future studies.

Furthermore, this study revealed significantly reduced odds of cohabiting with MAMM for children residing in the North-East geopolitical zone of Nigeria compared with those living in the North Central zone. However, it is harmful to a child living in South-East or South-South geopolitical zones to cohabit with MAMM compared to a child residing in North-Central geopolitical zones of Nigeria when other predictors are held constant. This finding agrees with a similar study [37], that under-five mortality is often high in the Northeast and Southeast of Nigeria, but does not agree with another study [51], especially when these childhood morbidities are treated independently. Finally, place of residence was a significant predictor of MAMM. The results from this study show that children in rural areas of Nigeria were at increased risk of cohabiting with MAMM when all other covariates were held constant. This finding did not agree with another study [36]

### Strengths and limitations of the study

The data set from the 2018 NDHS, which included merged contextual characteristics from the 2018 NHDR, was used in this study. Anaemia, malaria, and malnutrition are three objectively assessed (standard WHO measurement procedures) paediatric diseases combined for the first time in DHS data collection. So, to the best of the researcher's knowledge, this study is the first joint modelling of these diseases among children aged 6-59 months in Nigeria or anywhere else that is undertaken on the national scale. Similar studies have used self-reported assessment of other disease conditions, which might have introduced bias through under or over-reporting cases. Since this was a baseline study, it took a more simplified approach by developing a composite score that indicated the order in which the combination of these diseases occurred by the generally accepted definition of multimorbidity, as the cooccurrence of two or more diseases in an individual without reference to an index disease.

Additionally, the study realised that the inferential analysis strategy would not have been the best. However, as a baseline study, this will attract the interest of more research to better understand the determinants of multimorbidity in children in LMIC. Furthermore, to account for the effects of both individual and contextual variables on multimorbidity among children in Nigeria, this study recognised the hierarchical characteristics of the data set employed and used multilevel mixed effect ordinal logistic regression modelling.

Also, the data set came from a nationally representative survey with abundant evidence of hierarchy. Yet, most previous studies did not account for the multilevel structure or use the proper statistical techniques. This study applied multilevel methods to account for individual, community, and state variations.

However, this study is not without some limitations. First, the survey being cross-sectional, the study could only examine the associations between variables. Therefore causality could not be ascertained [37,52,53]. Though longitudinal research on children could be challenging in LMICs, they are needed to explain these predictors' significance and determine causal effects over time. Secondly, the study assumed that the three diseases were of equal importance or severity (e.g., having malaria was the same as having anaemia or malnutrition). However, the fulfilments of the proportional odds assumptions have to some extent, given credence to this. Thirdly, only Nigeria is the subject of this study, which limits its relevance or capacity to be generalised to other SSA countries.

Furthermore, it is difficult to directly compare our findings to those of past research due to the methodological differences in the study setting, disease scope, and demography. Furthermore, part of the findings was that children who have contracted malaria fever are more likely to be anaemic. In contrast, children being anaemic is harmful to being both malaria positive and poorly nourished and being poorly nourished is harmful to being anaemic. Given this, endogeneity biases may have resulted in the study. However, in multimorbidity, attempts were made in the first instance to categorize the three outcomes into eight independent classes (Section 4.3.3) from where the 'none of the diseases', 'one disease only', and 'two or more diseases' classes were derived. Therefore, if endogeneity had been ignored in this investigation, we might have certified some predictors significant when they could have just as quickly been due to chance [54]. Possibly the wrong assumptions were drawn. This finding could be a subject of future investigation. In addition, only the individual level that weighting was considered. These are suitable for future investigation.

## Policy and study implications

### Policy Implications

This study found that, at the time of the survey, about one in every two (48.3%) Nigerian children aged 6-59 months cohabits with two or more diseases of anaemia, malaria, and malnutrition. This result is alarming on a national scale and will likely be the case for children in Sub-Saharan Africa (SSA). It requires an urgent policy and a coordinated approach to check this from further escalation. This result shows that the situation of multimorbidity among children aged 6-59 months in Nigeria could be worst if nothing is done to reduce its prevalence and therefore becomes impossible for Nigeria to attain the SDG-3 by 2030. Some areas of urgent attention are:

1. The regional and state spatial maps reveal that some geopolitical regions and the states associated with them have high rates of diseases. For example, the North-East and North-West geopolitical zones have shared links to a high rate of insecurities for more than a decade, leaving the people, especially women and children, homeless (living in internally displaced person's camps). These people cannot carry out their farming activities, so they are destitute of good foods and drinking water. These spatial map descriptions of the prevalence of MAMM could be used by decision-makers to quickly target development efforts for health care, food security and poverty alleviation interventions in these areas of the high prevalence of MAMM. Ayala & Meier's article acknowledges the critical connections between the normative characteristics of the right to health and the four main components of food security (access, availability, stability, and utilisation). It views nutrition security as influencing public health (i.e., availability, accessibility, affordability, and quality) [55]. Hence, success-driven efforts to end the aged-long security issues in these regions so that people already displaced can return to their homes to farm again.
2. This study recognises some child-related risk factors of children cohabiting with MAMM. Firstly, male children are more likely than female children to contract MAMM. In the past (pre-millennium development goals era), gender inequalities were seen to be at a disadvantage of female children [56]. Still, in recent studies [53], and across the various disease spectrums in this study, female children were at an advantage. To address the disparities between genders in childhood MAMM in Nigeria community mobilisation against gender-based prejudice and gender-sensitive policies are needed. Secondly, children between the ages of one and three years were more likely to cohabitate with MAMM. These children are transitioning from being weaned from breastfeeding into supplementary feedings. Most children at this stage would not like to eat any other food introduced rather than continue with breast milk while the mother is unwilling to give. So, nutrient-fortified meals are given to children in this age group that could help sustain the immunity from their mother's breast milk until their natural immunity is built up at a later age. Thirdly, children born small (low birthweight) were at greater risk of MAMM when compared with children born larger birthweight. There are three primary reasons a child may be born with low birth weight (LBW). It could be due to genetics because the parents are small, or intrauterine growth restriction (IUGR) [57], or because the baby is born pre-term (before 37 weeks of pregnancy) [58]. To detect issues with foetal growth antenatal care is crucial and strongly encouraged for every pregnant woman. The ongoing reforms in Nigeria's health sector aim to dramatically alter the way that healthcare is delivered throughout Nigeria by increasing the accessibility, cost, quality, and availability of healthcare services via promoting private sector investment and engagement [59], making primary health care services accessible to the grassroots, especially to women and children should be the foremost priority. In addition, this study found that children with preceding birth order of 8-24 months are more likely to cohabit with two or more of anaemia, malaria, and malnutrition. As the interval age increases, the tendency to have MAMM in children drops significantly. Therefore, it is strongly advised that would-be mothers should adequately space their children above 24 months. This recommendation is because mothers who had their next child waiting for at least two years would have recovered most of the body's nutrients and blood loss during the first pregnancy and breastfeeding [60]. Finally, on child-related variables, children who were dewormed in the six months before the survey and those who had fever two weeks before the survey took place were more likely to be diagnosed with MAMM. It was recently reported that the Nigerian government had budgeted in 2021 the sum of N142.3 billion to feed about 10 million, deworming 7 million primary-level pupils in 35 states and the FCT [61]. However, this study has revealed that an estimated 38% of children of pre-school age who had not been dewormed are exposed to MAMM. So, a deworming program that will include children under-five should also be initiated. On the other hand, viral and bacterial infections are major causes of non-malaria fever among a significant number of children in Nigeria [62,63]. Therefore, as part of health education at antenatal clinics, it is crucial to stress the importance of personal and environmental cleanliness initiatives. In addition, children's immunisation against infectious diseases should be monitored adequately [64,65].
3. Regarding parental-related characteristics, maternal education, religion, anaemia status, and body mass index were significant predictors of MAMM among children aged 6-59 months in Nigeria. This finding shows that as mothers' education status increases, the probability of their children cohabiting with MAMM drops. The reason is that these mothers were likely to have more robust knowledge about health care to safeguard their children better and handle these illnesses [51]. Given this, girl-child education should be encouraged, especially in the northern part of the country, where MAMM is highly prevalent. Also, the findings show that children born to Muslim mothers were significantly more likely to contract MAMM compared to children of mothers of Catholic origin. One of the most significant influences on behaviour, particularly the pursuit of health among Nigerians, is religion [66]. Some religious organisations forbid their followers to participate in some health interventions. Even without promoting any particular religion, a pro-religion public policy can improve the health of the populace, including that of children [67]. Therefore, religious leaders should be allowed to participate in reaching evidence-based decisions to implement health intervention strategies that may directly affect their members.
4. Furthermore, children whose mothers are anaemic were 60% more likely to be anaemic than children whose mothers are not. The mechanism through which maternal anaemia status can affect children 6-59 months of age is complex. More importantly are the causes of maternal anaemia, especially during pregnancy which include poor diet (deficiencies in iron, folic acid, and vitamins), viral infections like malaria, and untreated hereditary haemoglobin abnormalities [40,68]. Moreover, the anaemic mother has no proper immunity to pass on to breastfeeding children and, as such, could easily be exposed to MAMM. Therefore, public health initiatives to lower childhood MAMM should pay more attention to reducing poor nutrition and other infections among pregnant, nursing mothers and children still breastfeeding. For instance, between 2013 and 2025, the Nigerian National Policy on Food and Nutrition has as one of its goals the reduction of maternal anaemia during pregnancy by 27% [21,69]. Therefore, there should be political commitment to make this happen. The findings also revealed that children whose mothers are overweight or obese were more protected from contracting MAMM. As earlier asserted, overweight mothers in developing countries are more likely to come from wealthy homes where eating good food is no problem. Though being overweight and obese have their health challenges. However, the high rate of poverty among families in Nigeria should be addressed. This provision will enable families able to afford good nutrition sources.
5. From the group of household-related predictors, only household wealth quintiles had significant effects on childhood MAMM. These results have further displayed the existence of two major social-economic classes concerning the health status of the children (The poor and the rich). To address this situation, social security that will take care of the immediate necessities of life (food, shelter, and health) for poor households in Nigeria should be in place.
6. In the community-related characteristics, children living in communities where the proportions of wealth status and distance to a health facility are no big problem are high and were significantly protective of MAMM compared to the low categories. Just as in the individual household characteristics, an increased community wealth level can bring about the same 'ripple' effects on childhood MAMM. The community with high wealth status would usually attract good facilities like clean water, good roads, and health facilities. Therefore, programs that bring many people out of poverty should be initiated and pursued vigorously. Children living in a community where the proportion of respondents who say that 'distance to the nearest health facility is no big problem' is high has demonstrated a reduced odd of contracting MAMM. This report implies that children accessing health facilities promptly are less likely to contract MAMM. Therefore, public health measures to lower childhood MAMM should pay more attention to increasing access to health centres through expanding the primary health care system.
7. Finally, the results show that the area-related variables, region, and place of residence were statistically significant predictors of MAMM. Children living in North-East, South-East, South-South, and South-West were more likely to cohabit with two or more of anaemia, malaria, and malnutrition than children living in North Central geopolitical zones. Efforts should be made to end the insecurity in the North-East and environmental degradation going on in South-East and South-South due to oil spillage that may have affected quite several things such as drinking water and farm produce. Children from the South-West have consistently increased odds of contracting any of the three diseases and the MAMM. This region seems more economically viable and educated, yet the children have a higher probability of cohabiting with two or more of these diseases. Finally, children residing in rural areas were more likely to be exposed to MAMM. Therefore, the public health approach should include making health care facilities closer to the people in rural areas. More importantly, health workers posted to these rural areas ensured they were well renumerated and monitored to dwell among the people.

### Study implications

Studies on multimorbidity in the adult population worldwide, especially in High-Income Countries (HICs), are well established. However, very few studies have been carried out on the children population, especially among children under-five years. Moreover, most of the researchers involved in multimorbidity studies are based in HICs. Therefore, they have not considered studying the cooccurrence of diseases in children because, with the efficient healthcare system that caters to the children population's health needs, multimorbidity in young children is not of many burdens. Conversely, in LMICs with a weak health care system accompanied by poverty, the health needs of children are often not taken as a priority, coupled with a lack of adequate data and researchers to conduct research that would help policymakers for informed decisions to provide integrated care for those co-inhabiting with multiple diseases makes the situation even scary.

This study is the first to examine the multiple overlaps of MAMM in Nigeria, for which nationally representative data have been made available for the first time; it serves as a baseline for other studies to build on. In addition, a recent study carried out by the Academy of Medical Sciences [70] has itemised the priority research needed to handle the global prevalence of multimorbidity in the adult population, which researchers in developing countries can as well see how these priority needs could be modified and adopted to stop the growing burden of childhood multimorbidity. The areas of urgent concern include:

1. A proper definition of multimorbidity that will be adaptable for the study of coexisting diseases among children in developing countries
2. The research should draw stakeholders' attention to the growing trends and prevalence of multimorbidity among children under-five years in developing countries.
3. Further research with appropriate statistical techniques to identify and describe these coexisting disease clusters among children.
4. Research verifying the individual's behavioural, biological, demographic, and environmental determinants of multimorbidity clustering to allow for direct comparison of studies.
5. To establish a well-coordinated cost-effective care approach beyond caring for individual diseases.

Additionally, funding bodies should show more interest in funding research on childhood multimorbidity in developing countries. This current study initiative has a baseline approach. Several alternative statistical analysis methods could be adopted to understand multimorbidity in children better while considering the model's requirement for parsimony (interpreting the results) from the available data. These methods include multivariate joint logistic regression, multinomial logistic regression, latent variables determination, and simultaneous equation model [53,71–73]. Even though this study used a more traditional frequentist method, it looked at the spatial map distribution of prevalence of MAMM across the six geopolitical zones, the states, and the FCT. Since all predictors were categorised, some metric covariates could have had non-linear effects on MAMM if employed as-is [53,74,75]; a future study might use a strictly Bayesian approach with random effect components. The method is expedient for comparing models from the standpoint of Classical and Bayesian approaches and offers a suitable model for analysis of MAMM. Furthermore, although longitudinal studies on children in LMICs may be difficult, they are required to demonstrate the importance of the predictors of MAMM and identify causal relationships across time. When longitudinal data becomes available for these diseases, a more sophisticated modelling approach on the connection between anaemia, malaria, and malnutrition can be developed to enhance integrated care for children.

## Conclusion

The three adverse health outcomes (anaemia, malaria, and malnutrition) clusters significantly contribute to Nigeria's child mortality. This study's objectives include determining the prevalence across states and regions in Nigeria. According to this study, two or more diseases are present in almost one-half of Nigerian children aged 6 to 59 months. This is worrying, an indication that health inequalities cut across the individual and contextual characteristics of children aged 6-59 months in Nigeria. Therefore, it requires urgent response through creating and executing good policies to address these situations if SDG-3 must be realized in Nigeria. In addition, the results have demonstrated the need for clinicians and health care providers to evolve integrated care models suitable for managing and treating children cohabiting with multiple diseases (especially, anaemia, malaria, and malnutrition). There is urgent need of paradigm shift in the training curriculum of medical school from the clinical guidelines of treating for single disease to include handling clusters of diseases especially among children in LMICs.

## Data Availability

The data underlying these findings are publicly available at the Demographic and Health Survey (DHS) https://dhsprogram.com/Data/ and the National Human Development Report (2018) of the United Nations Development Program UNDP-Nigeria. https://hdr.undp.org/content/national-human-development-report-2018-nigeria

https://hdr.undp.org/content/national-human-development-report-2018-nigeria

